# Phase angle is related to physical function and quality of life in preoperative patients with lumbar spinal stenosis

**DOI:** 10.1101/2022.11.02.22281869

**Authors:** Ryota Otsubo, Ryuki Hashida, Kenta Murotani, Sohei Iwanaga, Keisuke Hirota, Syunji Koya, Yuya Tsukada, Yuta Ogata, Kimiaki Yokosuka, Tatsuhiro Yoshida, Ichiro Nakae, Takuma Fudo, Shinji Morito, Takahiro Shimazaki, Shingo Shimogawa, Reika Yamaryo, Mai Ito, Kei Yamada, Kimiaki Sato, Hiroo Matsuse, Naoto Shiba, Koji Hiraoka

## Abstract

**Purpose:** Lumbar spinal stenosis (LSS) causes various symptoms, which can interfere with daily life and Quality of Life (QOL). Evaluating both patients’ physical function and QOL and helping them to improve is the focus of rehabilitation. Phase angle (PhA) assessment has been widely used to measure body composition, and it has been reported to reflect physical function and QOL. The purpose of this study was to investigate the relationship between PhA and physical function, physical activity, and QOL in patients with LSS.

**Methods:** PhA, handgrip strength, walking speed, timed up and go test (TUG), Life Space Assessment (LSA), Prognostic Nutritional Index (PNI), Japanese Orthopaedic Association back pain evaluation questionnaire (JOABPEQ), and EQ-5D were assessed. Multiple regression analysis was conducted to investigate the relationship of PhA to physical function, physical activity, and QOL using.

**Results:** Statistical analysis was performed on 104 patients with LSS. The results of multiple regression analysis of PhA adjusted for age, gender, and BMI (Model 1) and PhA adjusted for age, gender, BMI, and PNI (Model 2) compared with physical function and physical activity showed significant correlations respectively (P<0.05) in handgrip strength, walking speed, TUG, and LSA. In QOL assessment, both models showed a significant correlation (P<0.05) with lumbar function in JOABPEQ and a significant trend (P<0.1) in EQ-5D.

**Conclusion:** PhA in LSS patients in this study was associated with physical function and QOL, and may be a useful new tool for clinical evaluation in preoperative LSS patients.

## Introduction

Lumbar spinal stenosis (LSS) is a disease caused by narrowing of the spinal canal, impedes the nerves and blood vessels traveling through the lumbar spine. LSS causes pain in the lower back, buttocks, and lower extremities, which can interfere with daily life [1]. In addition, intermittent claudication is one of the most common symptoms of LSS, severely limiting the patient’s mobility. It has been reported that LSS with impaired activities of daily living may lead to psychosocial problems such as depression and isolation due to symptoms such as intermittent claudication, back pain, numbness, and muscle weakness in the lower limbs, leading to a decrease in quality of life (QOL) [2]. Improving QOL is one of the important goals of LSS treatment [3], and assessing QOL is also important in physical therapy.

QOL is a term that originated from the World Health Organization’s (WHO) 1947 Charter on Health, which defines health as not only the absence of disease, but also a state of physical, psychological, and social satisfaction. The evaluation of QOL is broadly classified into three categories: health-related QOL, non-health-related QOL and psychological QOL including wellbeing, happiness, and satisfaction with life. Of these, health-related QOL has been roughly divided into two scales: a disease-specific scale that evaluates specific symptoms of disease, and a comprehensive scale that evaluates general conditions regardless of the presence or absence of disease. The Japanese Orthopaedic Association Back Pain Evaluation Questionnaire (JOABPEQ) is a patient-administered scale that includes a lumbar spine disease-specific QOL assessment that can evaluate not only pain but also functional disability, gait dysfunction, social disadvantage, and psychological disability due to back pain [3]. The EuroQOL 5 dimensions 3-level (EQ-5D) is a scale that assesses a comprehensive health-related quality of life score that evaluates mobility, self-care, activity, pain, and depression on three levels [4].

In recent years, phase angle (PhA) assessment has been widely used to noninvasively measure body composition, and has been reported as a measure of skeletal muscle quality [5]. A high value of PhA reflects higher cellularity, cell membrane integrity and better cell function, while a low value reflects cell loss and reduced cell membrane integrity [6]. PhA has been reported to correlate with physical functions such as muscle strength, balance, and walking speed in both community-dwelling elderly and cancer patients [7, 8]. In maintenance hemodialysis patients, a correlation with health-related QOL has been reported [9, 10]. Therefore, PhA reflects physical function and QOL, and is considered to be a useful tool for understanding patients’ condition in clinical practice. However, although PhA is a non-invasive and simple method of measuring body composition, there have been few reports using PhA in orthopedic diseases, and its relationship with physical function and QOL has not yet been fully investigated. LSS is often associated with pain and numbness, which may make it difficult to assess physical functions such as muscle strength, balance function, and walking speed. In addition, physical function, activities of daily living, and QOL are interrelated, and PhA, which can measure them non-invasively, may be useful for indirect assessment of physical function and QOL in patients who are difficult to assess in practice.

The purpose of this study was to investigate the relationship between PhA, physical function, and QOL in patients with LSS, an orthopedic disease.

## Method

### Ethics

The study protocol conformed to the ethics guidelines of the Declaration of Helsinki, as reflected in prior approval given by the institutional review board of University Hospital (approval ID: 22017). This study was a retrospective study, thus we published documents of informed consent forms including the title of the study, subject, and aim on the web. Informed consent from patients was obtained using an opt-out approach.

### Patients

This cross-sectional study was conducted on 162 patients admitted for surgery for LSS in the orthopedic ward of our hospital between May 2020 and April 2021. Data collection was conducted in May 2022, and no personally identifiable participant information was accessed after data collection. The eligibility criteria for this study were patients who were admitted for surgery for lumbar spinal canal stenosis and were at least 60 years of age. The exclusion criteria were pyogenic spondylitis, metastatic bone tumor, history of stroke, difficulty in standing due to hemiplegia, difficulty in measuring body composition, insertion of a pacemaker, and inability to answer the questionnaire. The procedures for various examinations and evaluations were as follows: PhA was measured before admission, and physical function and QOL evaluations were performed before surgery.

### Phase angle (PhA)

PhA was measured using bioelectrical impedance analysis (BIA). An Inbody 770 BIA unit (Inbody Co., Ltd., Seoul, Korea) was used for the measurements. The BIA measures body composition by passing a weak alternating current through the body and taking the impedance or the difference between each tissue. Impedance is a general term for resistance components that interfere with the current, and impedance can be divided into resistance (R) and reactance (Xc). PhA is the value of arctangent, which is Xc divided by R, and is expressed by the following equation [6].

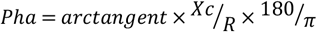

### Life Space Assessment (LSA)

The LSA is an index that assesses mobility in the spatial extent of an individual’s daily life [11]. This consists of the range of living space, frequency, presence of assistive devices, and level of independence, and is a questionnaire measure that investigates the range of activities in the living space over the past month. The living space from the bedroom at home to outside the town is divided into five levels, and the total score is calculated by multiplying the score of each level by the frequency and the degree of independence. The full score is 120, and a higher score means a higher level of activity. The LSA has been used to assess mobility and daily activities of living in a variety of conditions, including community-dwelling elderly and stroke patients [12, 13].

### Hand-grip strength

Grip strength was measured using a Smedley-type grip strength meter (Takei Kiki Kogyo, T.K.K. 5101) [14]. The subject was instructed, “After the signal, grip the grip strength meter as strongly as possible. Also, please hold the grip strength meter for 3 seconds.” The right hand was measured twice, and the maximum value was adopted. Grip strength is used to measure voluntary muscle function as an indicator of muscle strength in patients with LSS [15].

### Walking speed

A 10-meter walk test was conducted to calculate the walking speed. In the 10-meter walk test, two marks were placed on the floor so that the distance was 10 meters in a straight line, and the time it took from the leading foot passing the first mark to the moment it stepped on or over the second mark was measured with a stopwatch. To make sure that the subject started 1-meter before the first mark and passed the second mark by 1-meter, the subject was not told about the marks, but was shown the direction and instructed, “Walk straight ahead at your normal walking speed.” [16].

### Timed Up and Go test (TUG)

For the TUG, the subject was seated, and then asked “Please get up from the chair, walk as fast as you can, walk around the cone 3 meters ahead, and sit on the chair again. You may turn in either direction.” The TUG was measured with a stopwatch from the time the subject’s body began to move to the time they were seated on the chair again. A chair with a height of 42.0 cm and an elbow rest was used [17].

### Prognostic nutritional index (PNI)

PNI is also a simple and reliable assessment of nutritional status [18]. Consequently, PNI was also assessed in this study because it is predicted to be a confounder with nutrition. PNI is a prognostic score that is calculated by reflecting both the inflammatory and nutritional status of the patient. In the current study, it was calculated with the following formula using the preoperative blood test values.

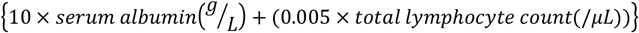

PNI has been reported to be a useful marker of malnutrition [19]. It has also been reported that low preoperative PNI is indicative of low nutritional status and is associated with postoperative complications in spinal diseases [20, 21].

### Japanese Orthopaedic Association Back Pain Evaluation Questionnaire (JOABPEQ)

The JOABPEQ is a back pain disease-specific QOL assessment that combines the Roland-Morris Disability Questionnaire (RDQ) and the Short-Form Health Survey (SF-36) with the Visual Analog Scale (VAS). The assessment consists of 25 items corresponding to five subscales: low back pain (4 items), lumbar function (6 items), walking ability (5 items), social life function (3 items), and mental health (7 items). The score for each subscale ranges from 0-100, with higher scores indicating better conditions [22].

### EuroQOL 5 dimensions 3-level (EQ-5D)

The EQ-5D is a comprehensive quality of life scale that can be calculated by answering five questions in three levels: “degree of mobility,” “personal care,” “usual activities,” “pain/discomfort,” and “anxiety/stress”. It has been used to assess health-related quality of life in middle-aged and elderly people living at home and for various diseases, including orthopedic diseases [23, 24].

### Statistical analysis

JMP version 15.0 was used for statistical analysis, and the significance level was set at 5%. As a statistical treatment, the association between PhA and each measurement item (age, gender, BMI, PNI, handgrip strength, walking speed, TUG, LSA, each sub-item of JOABPEQ, and EQ-5D) was investigated using Spearman’s rank correlation coefficient. The PhA has been influenced by three main factors: age, gender, and BMI [6], and has also been reported to be a potentially useful marker of malnutrition [19]. In the present study, we focused on the relationship between PhA, physical function, and quality of life. For this purpose, we created two models, one without adjustment by PNI (Model 1) and the other with adjustment (Model 2). In each model, we investigated the relationship between PhA and physical function (grip strength, walking speed, TUG), physical activity (LSA), and QOL (JOABPEQ sub-items, EQ-5D) using multiple regression analysis with PhA as the objective variable. Grip strength, TUG, and 10-meter walk tests are easy and practical tools to assess patients’ physical abilities, and these physical assessments have been used in patients with LSS in clinical settings [15, 25, 26]. Thus, we applied these items in this study.

## Result

### Patients

Of the 162 patients who met the eligibility criteria for this study, 104 patients were analyzed after removing the excluded cases (Figure 1). The basic attributes of the patients are shown in Table 1.

**Figure 1.**
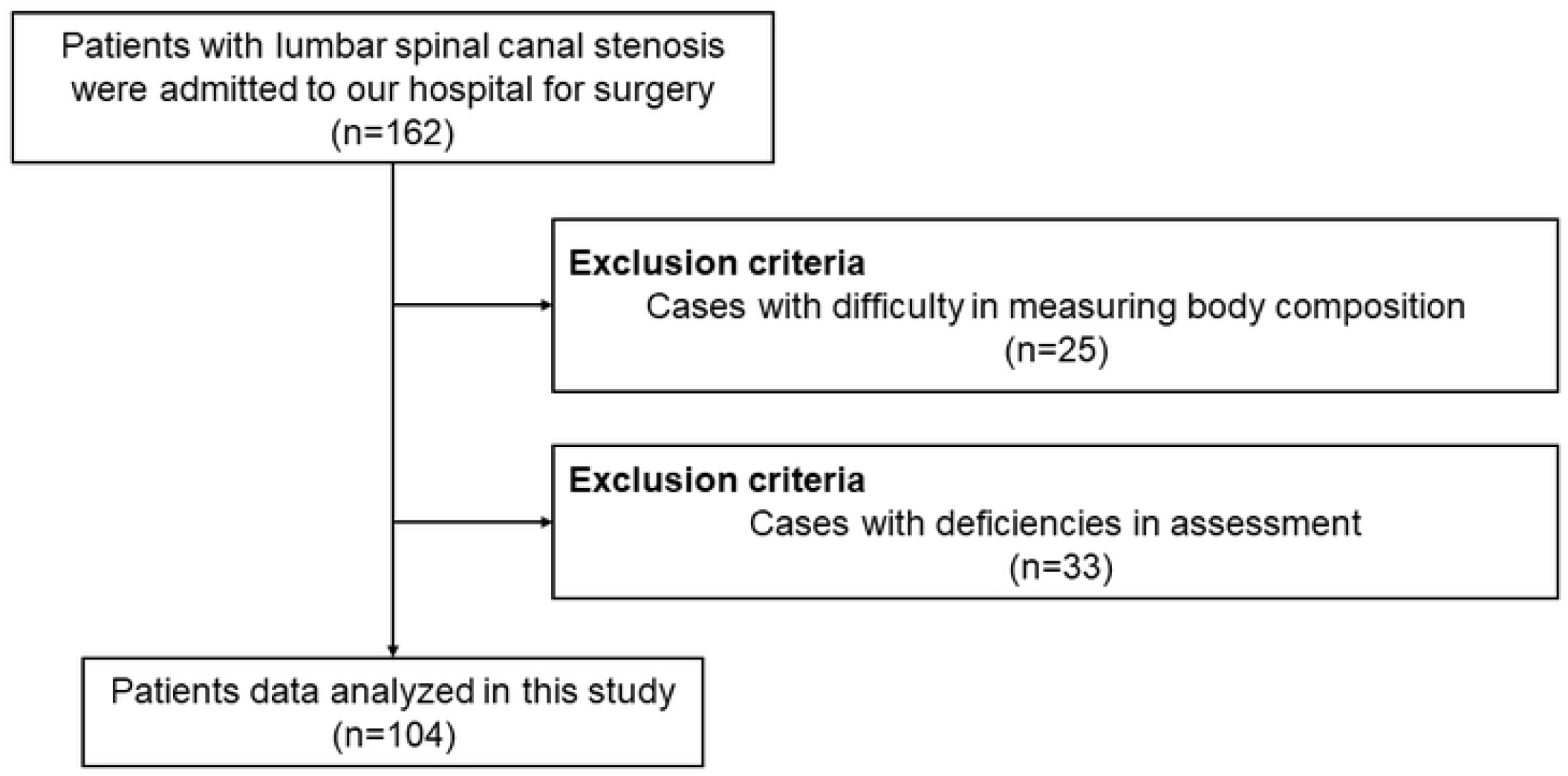
Diagram of patient’s inclusion and exclusion criteria in this study

**Table 1.**
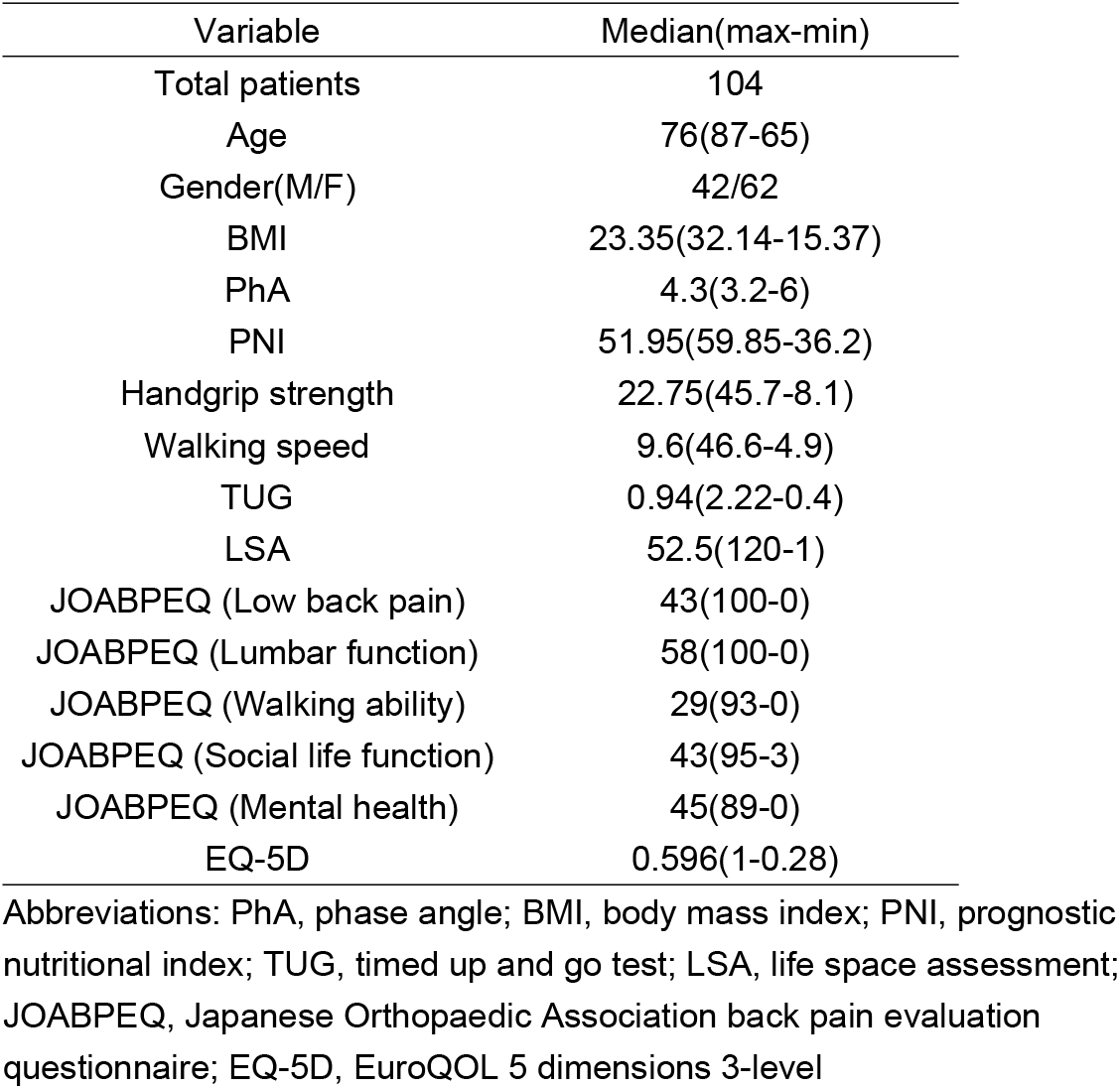
Characteristics

| Variable | Median(max-min) |
| --- | --- |
| Total patients | 104 |
| Age | 76(87-65) |
| Gender(M/F) | 42/62 |
| BMI | 23.35(32.14-15.37) |
| PhA | 4.3(3.2-6) |
| PNI | 51.95(59.85-36.2) |
| Handgrip strength | 22.75(45.7-8.1) |
| Walking speed | 9.6(46.6-4.9) |
| TUG | 0.94(2.22-0.4) |
| LSA | 52.5(120-1) |
| JOABPEQ (Low back pain) | 43(100-0) |
| JOABPEQ (Lumbar function) | 58(100-0) |
| JOABPEQ (Walking ability) | 29(93-0) |
| JOABPEQ (Social life function) | 43(95-3) |
| JOABPEQ (Mental health) | 45(89-0) |
| EQ-5D | 0.596(1-0.28) |

Abbreviations: PhA, phase angle; BMI, body mass index; PNI, prognostic nutritional index; TUG, timed up and go test; LSA, life space assessment; JOABPEQ, Japanese Orthopaedic Association back pain evaluation questionnaire; EQ-5D, EuroQOL 5 dimensions 3-level

### Correlation between PhA and each measurement item

The results of Spearman’s rank correlation coefficients between PhA and each measurement item are shown in Table 2. PhA showed a positive and negative correlation with the following items. PhA was positively correlated with PNI, handgrip strength, walking speed, LSA. PhA was negatively correlated with age and TUG. PhA showed a trend with BMI.

**Table 2.** Spearman’s rank correlation coefficient (PhA vs each measurement item)

| Variable | vs Variable | $\rho$ | P-value |
| --- | --- | --- | --- |
| Phase Angle | Age | -0.443 | <.0001* |
|  | BMI | 0.182 | 0.0646† |
|  | PNI | 0.193 | 0.0499* |
|  | Handgrip strength | 0.561 | <.0001* |
|  | Walking speed | 0.476 | <.0001* |
|  | TUG | -0.498 | <.0001* |
|  | LSA | 0.475 | <.0001* |
|  | JOABPEQ (Low back pain) | -0.003 | 0.9720 |
|  | JOABPEQ (Lumbar function) | 0.299 | 0.0021* |
|  | JOABPEQ (Walking ability) | 0.108 | 0.2764 |
|  | JOABPEQ (Social life function) | 0.233 | 0.0175* |
|  | JOABPEQ (Mental health) | 0.277 | 0.0044* |
|  | EQ-5D | 0.262 | 0.0071* |

Abbreviations: PhA, phase angle; BMI, body mass index; PNI, prognostic nutritional index; TUG, timed up and go test; LSA, life space assessment; JOABPEQ, Japanese Orthopaedic Association back pain evaluation questionnaire; EQ-5D, EuroQOL 5 dimensions 3-level
※P-value<0.1†、P-value<0.05\*

PhA and QOL assessment (EQ-5D and JOABPEQ items) showed a positive correlation in Lumbar function, social life function, and mental health, which are sub-items of JOABPEQ, and also a positive correlation in EQ-5D.

PhA was negatively correlated with age and showed a trend with BMI; PhA was positively correlated with LSA, handgrip strength, and walking speed in physical activity and physical function assessment, and negatively correlated with TUG test. PhA and PNI showed a positive correlation. PhA and QOL assessment (EQ-5D and JOABPEQ items) showed a positive correlation in Lumbar function, social life function, and mental health, which are sub-items of JOABPEQ, and also a positive correlation in EQ-5D.

### Multiple regression analysis of confounding-adjusted PhA, physical function, and quality of life

The results of multiple regression analysis between both PhA adjusted for age, gender, and BMI (Model 1) and PhA adjusted for age, gender, BMI, and PNI (Model 2) and physical function and physical activity are shown in Table 3. Moreover, the results of multiple regression analysis between both PhA and QOL assessment adjusted by Model 1, 2 are shown in Table 4. In the evaluation of physical function and physical activity, significant correlations were found for all of the handgrip strength, walking speed, TUG, and LSA measured in this study, and in QOL, significant correlations were found with the lumbar function of JOABPEQ in both Model 1 and Model 2, and a significant trend was found in EQ-5D.

**Table 3.** Multiple regression analysis (PhA vs Physical activity and Physical function)

|  | Outcome | Model | Estimated value | 95%CI | P-value |
| --- | --- | --- | --- | --- | --- |
| Physical activity | LSA | 1 | 19.777 | 8.444 – 31.110 | 0.0008* |
|  |  | 2 | 21.063 | 9.581 – 32.544 | 0.0004* |
| Physical function | Handgrip strength | 1 | 3.578 | 1.126 – 6.031 | 0.0047* |
|  |  | 2 | 3.447 | 0.947 – 5.947 | 0.0074* |
|  | Walking speed | 1 | 0.293 | 0.145 – 0.440 | 0.0002* |
|  |  | 2 | 0.304 | 0.154 – 0.454 | 0.0001* |
|  | TUG | 1 | -5.460 | -8.665 – -2.254 | 0.0010* |
|  |  | 2 | -5.353 | -8.624 – -2.082 | 0.0016* |

Abbreviations: PhA, phase angle; CI, confidence interval; LSA, life space assessment; TUG, timed up and go test
※Model 1: Adjusted for age, gender, and BMI
※Model 2: Adjusted for age, gender, BMI, and PNI
※P-value<0.1†,P-value<0.05\*

**Table 4.** Multiple regression analysis (PhA vs QOL)

| Outcome |  | Model | Estimated value | 95%CI | P-value |
| --- | --- | --- | --- | --- | --- |
| Disease-specific QOL | JOABPEQ (Low back pain) | 1 | 0.080 | -15.479 – 15.638 | 0.9919 |
|  |  | 2 | 1.100 | -14.748 – 16.947 | 0.8908 |
|  | JOABPEQ (Lumbar function) | 1 | 16.924 | 4.641 – 29.207 | 0.0074* |
|  |  | 2 | 16.169 | 3.654 – 28.684 | 0.0119* |
|  | JOABPEQ (Walking ability) | 1 | 4.401 | -6.642 – 15.444 | 0.4310 |
|  |  | 2 | 5.924 | -5.220 – 17.069 | 0.2941 |
|  | JOABPEQ (Social life function) | 1 | 5.923 | -2.397 – 14.242 | 0.1609 |
|  |  | 2 | 7.061 | -1.337 – 15.459 | 0.0984† |
|  | JOABPEQ (Mental health) | 1 | 6.388 | -1.113 – 13.889 | 0.0942† |
|  |  | 2 | 5.884 | -1.755 – 13.524 | 0.1296 |
| Comprehensive QOL | EQ-5D | 1 | 0.044 | -0.004 – 0.093 | 0.0737† |
|  |  | 2 | 0.043 | -0.006 – 0.092 | 0.0897† |

Abbreviations: PhA, phase angle; QOL, quality of life; CI, confidence interval; JOABPEQ, Japanese Orthopaedic Association back pain evaluation questionnaire; EQ-5D, EuroQOL 5 dimensions 3-level
※Model 1: Adjusted for age, gender, and BMI
※Model 2: Adjusted for age, gender, BMI, and PNI
※P-value<0.1†, P-value<0.05

## Discussion

In this study, we investigated the relationship between PhA and physical function, physical activity, and QOL in LSS patients admitted for surgery. A significant correlation was found between PhA and physical function and physical activity, and PhA was correlated with the sub-items of JOABPEQ, a disease-specific QOL assessment, and EQ-5D, a comprehensive QOL assessment. Furthermore, in multiple regression analysis, both Model 1 and Model 2 showed a correlation with lumbar function, a sub-item of the JOABPEQ. PhA may be considered a useful tool for clinical and perioperative evaluation because it reflects not only physical function but also QOL (Figure2).

**Figure 2.**
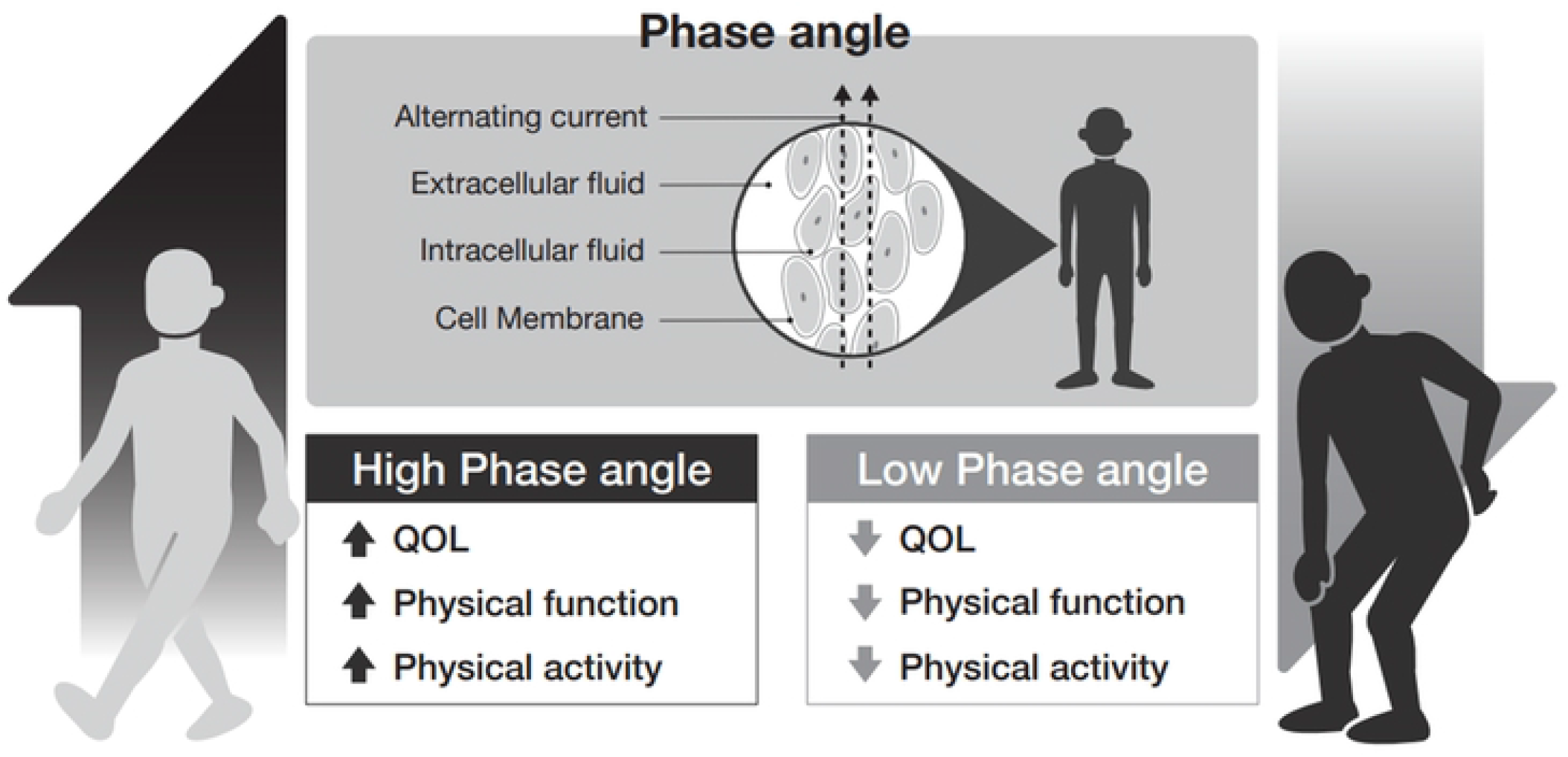
Graphical abstract

Summary of this study. The phase angle using bioelectrical impedance analysis was associated with quality of life, physical function, and physical activity in patients with lumbar spinal stenosis.

### The relationship between PhA, physical function, and physical activity

In the present study, significant positive correlations were found between PhA of LSS patients and handgrip strength and walking speed, and significant negative correlations were found with the TUG test. Yamada et al. reported that PhA was significantly correlated with lower limb muscle strength in community-dwelling elderly people [27]. Tanaka et al. in their study of the locomotive syndrome (LS) and PhA in community-dwelling elderly, found that PhA was significantly correlated with the TUG test, and furthermore, PhA was an independent factor for LS [28]. In the present study conducted on patients with LSS as well as in previous studies on other diseases, PhA was significantly higher in patients with higher physical function.

Furthermore, in the present study, a significant positive correlation was also found between LSA and PhA, which indicates physical activity. High physical activity is associated with high physical function, and a correlation between physical activity and PhA has been reported in other studies [18]. However, the mechanisms of physical activity, physical function, and PhA are unknown. Based on the principles of PhA measurement, it is likely that the reason for the results of the present study is that the decrease in the amount of fat, which has a high electrical resistance, due to physical activity, decreases the resistance, and the increase in the size and mass of cells and the structural integrity of cell membranes increases the reactance (Xc), which in turn results in a higher PhA [6].

### About PhA and JOABPEQ

PhA was correlated with lumbar function on the JOABPEQ in both Model 1 and Model 2. The items of the JOABPEQ questioners for lumbar function included the following. “I sometimes ask for help when I do something because of my back pain,” “I try not to bend my back or kneel because of my back pain,” “I have difficulty getting up from a chair because of my back pain,” “I have difficulty turning over because of my back pain,” “I have difficulty putting on socks or stockings because of my back pain” and “Do you find it difficult to bend forward, kneel, or bend over because of your physical condition?”. These questions are related to the flexion, extension, and rotation of the trunk. For preoperative LSS patients with low back pain, trunk flexion, extension, and rotation movements should not be assessed because they are likely to induce pain. Therefore, PhA may be one of the indicators to evaluate lumbar function in patients with LSS because it can estimate lumbar function without actual movements.

### About PhA and Quality of Life

In the present study, PhA were significant correlations found between both Model 1 and Model 2 and the lumbar function sub-items of the JOABPEQ, a disease-specific QOL scale, as well as a trend was found in the EQ-5D, a comprehensive QOL scale. Machado et al. demonstrated that PhA is associated with not only physical function but also QOL in patients with idiopathic pulmonary fibrosis [29]. Azar et al. reported in their study of dialysis patients that a clinically useful predictive tool because PhA reflects also physical function and QOL [10]. This is the first study to investigate the relationship between PhA and quality of life in LSS, with results similar to other previous studies. The JOABPEQ is a lumbar spine disease-specific quality of life measure and is a highly responsive outcome measure for assessing changes over time in patients with low back pain [22]. The EQ-5D, on the other hand, is a widely used comprehensive QOL assessment [30]. The study found that both disease-specific and comprehensive QOL was associated with PhA, suggesting that PhA and QOL may be related in patients with LSS. Few studies have investigated both QOL measures, and the results of this study seem to confirm the above findings. Moreover, as mentioned earlier, PhA also reflected the physical function (lumbar spine function) and QOL of LSS patients in this study. Evaluating both physical function and QOL of patients and helping them to improve is the focus of rehabilitation [31]. Therefore, PhA may be one useful tool in physical therapy in patients with LSS.

## Limitation

There are several limitations of this study that should be considered. First, it is a cross-sectional study, and the causal relationship is unclear because the broader relationship between disease and factors is not known due to the design. In addition, this was a single-center study with a small number of cases and no direct assessment of lumbar function. In this study, only preoperative patients were studied, and future studies of patients who are not suitable for surgery and postoperative patients should be examined. Moreover, we demonstrated that PhA is associated with QOL and physical function in patients with LSS in this study, however, we didn’t compare it with healthy subjects or other diseases. The extent of spinal canal narrowing correlates with the limitation of LSS-typical claudication. We didn’t investigate the radiological extent of spinal canal narrowing. We need to investigate the radiological extent of spinal canal narrowing in a future study.

## Conclusion

PhA was found to be associated with physical function and QOL in LSS patients. PhA may be a useful new tool for clinical assessment in preoperative LSS patients.

## Data Availability

Hashida, Ryuki Otsubo Ryota, 2022, "Phase angle is related to physical function and quality of life in preoperative patients with lumbar spinal stenosis", https://doi.org/10.7910/DVN/REBPZ4, Harvard Dataverse, V1

